# Epigenome-wide DNA methylation profiling of healthy COVID-19 recoverees reveals a unique signature in circulating immune cells

**DOI:** 10.1101/2021.07.05.21260014

**Authors:** Johanna Huoman, Shumaila Sayyab, Eirini Apostolou, Lovisa Karlsson, Lucas Porcile, Muhammad Rizwan, Sumit Sharma, Jyotirmoy Das, Anders Rosén, Maria Lerm

## Abstract

**Background:** Epigenetic alterations upon microbial challenge have been described as both a defence strategy and a result of pathogenic manipulation. While most COVID-19 studies focus on inflammatory and immune-mediated responses, little is known about epigenetic modifications in response to SARS-CoV-2 infection.

**Methods:** Epigenome-wide DNA methylation patterns from COVID-19 convalescents were compared to uninfected controls from before and after the pandemic. Peripheral blood mononuclear cell (PBMC) DNA was extracted from uninfected controls, COVID-19 convalescents and symptom-free individuals with SARS-CoV-2-specific T cell-responses, as well as from PBMCs stimulated *in vitro* with SARS-CoV-2. Subsequently, the Illumina MethylationEPIC 850K array was performed, and statistical/bioinformatic analyses comprised differential DNA methylation, pathway over-representation and module identification analyses.

**Results:** Differential DNA methylation patterns distinguished COVID-19 convalescents from uninfected controls, with similar results in an experimental SARS-CoV-2 infection model. A SARS-CoV-2-induced module was identified *in vivo*, comprising 66 genes of which six (*TP53, INS, HSPA4, SP1, ESR1* and *FAS*) were present in corresponding *in vitro* analyses. Over-representation analyses revealed involvement in Wnt, muscarinic acetylcholine receptor signalling and gonadotropin-releasing hormone receptor pathways. Furthermore, numerous differentially methylated and network genes from both settings interacted with the SARS-CoV-2 interactome.

**Conclusions:** Altered DNA methylation patterns of COVID-19 convalescents suggest recovery from mild-to-moderate SARS-CoV-2 infection leaves longstanding epigenetic traces. As *in vitro* SARS-CoV-2 infection corroborated *in vivo* exposure results, this indicates DNA methylation is involved in immune cell responses to challenge with this virus. Future studies should determine whether this reflects host-induced protective antiviral defence or targeted viral hijacking to evade host defence.

## INTRODUCTION

Since the end of 2019, the Corona virus disease 19 (COVID-19) pandemic has claimed lives of millions world-wide, highlighting the global challenges in detecting, monitoring, and treating novel viral infections. While efficacious vaccines are available at present, still a lot remains to be uncovered regarding underlying mechanisms of the interaction between the severe acute respiratory syndrome coronavirus 2 (SARS-CoV-2) virus and its host.

SARS-CoV-2 is a single stranded enveloped RNA virus belonging to the *Coronaviridae* family^1^, which similarly to other viruses hijacks host functions for its own advantage.^2-5^ Evidence suggest that epigenetic mechanisms, *i.e*. processes regulating transcriptional accessibility of genomes without altering the nucleic acid sequence, are involved in the hijacking process.^6, 7^ DNA methylation (DNAm) of CpG sites is considered to be the most stable epigenetic modification, as it ensures heritability throughout cell division, although it is at the same time highly dynamic in response to environmental stimuli.^8^ The malleability and flexibility of the DNA methylome decreases with increasing age^9^, and environmental factors such as smoking and nutrition may alter DNAm patterning in various cell types, including different immune cells.^10, 11^ This could have important implications in the course of COVID-19, as *e.g*. smokers, overweight and elderly patients are more susceptible to become severely ill if contracting SARS-CoV-2.^12, 13^ Furthermore, DNAm patterns may also become altered upon microbial^14^ or viral infection.^15-17^ In line with this, we have previously observed that immune cells of asymptomatic, tuberculosis-exposed individuals carry a lasting DNAm signature that is linked to protection against mycobacterial infection.^18-20^

A majority (40-80%) of individuals infected with SARS-CoV-2 show no or mild symptoms of COVID-19 and proceed into convalescence thereafter, while a smaller, but non-negligible, proportion of individuals show severe or life-threatening manifestations.^21, 22^ Tolerant immune responses have been observed in transcriptomic and immune profiling comparisons of asymptomatic and symptomatic COVID-19 patients^23^, and as epigenetic mechanisms regulate differentiation of *e.g*. T cells^8^, it is conceivable that epigenetic mechanisms may be implicated in combating SARS-CoV-2 infection. However, few studies have thus far addressed whether and how the epigenome is altered in subjects with a recent mild-to-moderate SARS-CoV-2 infection. In this study, we set out to examine epigenome-wide DNAm patterns in convalescent COVID-19 (CC19) subjects, after a mild-to-moderate disease course. Understanding how convalescent individuals have mounted an epigenetic response against new viruses such as SARS-CoV-2, for which no pre-existent immunity was present, may reveal how a functional defence strategy towards the virus is prepared, and guide development of novel diagnostic and preventive measures. Indeed, we found that differential DNAm patterns separated those who have not been infected with SARS-CoV-2 from those who have recovered from mild COVID-19, suggesting that epigenetic mechanisms are at play during SARS-CoV-2 infection. The observations could be replicated in *in vitro* experiments, further underpinning our findings.

## MATERIALS AND METHODS

### Study population

In this study, participants were enrolled between May 29^th^ and July 10^th^ 2020 during the first wave of the SARS-CoV-2 pandemic in Linköping, Sweden. Individuals who had recovered from and individuals who had not experienced COVID-19 were recruited after announcements with leaflets. Exclusion criteria were the existence of current active SARS-CoV-2 infection and/or other infectious disease symptoms, as well as being younger than 18 years. The study participants voluntarily entered the study in a consecutive manner. The study was conducted on blood and saliva samples from in total 38 individuals from three different groups (Figure 1); non-infected controls (Con, n=18), COVID-19 convalescents (CC19, n=14) and symptom-free individuals with SARS-CoV-2-specific T cell responses (SFT, n=6). Additionally, blood samples from anonymous healthy blood donors from the blood bank at Linköping University Hospital before 2020 were included as a separate group in the analyses (pre20, n=5), collected between 2014-2019 prior to the outbreak of the pandemic. CC19 participants presented with either mild or asymptomatic initial infection, and none was admitted to hospital. Cons were defined as neither having any positive circulating IgG-antibody or T cell responses to SARS-CoV-2, while CC19s were defined by the presence of SARS-CoV-2-specific IgG antibodies in plasma using suspension multiplex immunoassay (SMIA), some of which were positive for IgG in saliva, rapid test and in T cell responses as well. From the included individuals, the following information was retrieved using health questionnaires: self-reported COVID-19 symptoms (if applicable, one or several of the following: fever, headache, shortness of breath, loss of smell/taste, cough, fatigue, muscle pain, nausea, sinusitis/congestion), date of self-reported symptoms, weeks between symptoms and sampling, age, sex, smoking, weight, height, comorbidities as well as medications. The blood and saliva from the study participants was processed in a Biosafety level-2 facility. The present study is an exploratory pilot study on the effects of mild-to-moderate SARS-CoV-2 infection on DNA methylation patterns in PBMCs. The sample size could not be determined beforehand, as the SARS-CoV-2 infection rate in society was not known at the time of sample collection. Hence, all individuals fulfilling inclusion criteria, consenting to participation and providing both blood and saliva samples were included in the study. However, the sample size is similar to previous studies on the effect of BCG vaccination^18, 24^, where meaningful differences in DNA methylation upon tuberculosis infection were shown. Likewise, the belonging to the different groups described above was determined after performance of the DNA methylation analyses, and hence handling, extraction and experimental procedures performed on the samples were performed in a blinded fashion. For samples from the natural exposure cohort, all participants provided written informed consent, and the present study was approved by the Regional Ethics Committee for Human Research in Linköping (Dnr. 2019-0618). Regarding the anonymous blood samples used for *in vitro* experiments, informed consent was given by the healthy donors at the time of blood donation and the use of the donated blood for research purposes was guaranteed as per the guidelines of Regional Ethics Committee for Human Research in Linköping and the Helsinki Declaration.

**Figure 1.**
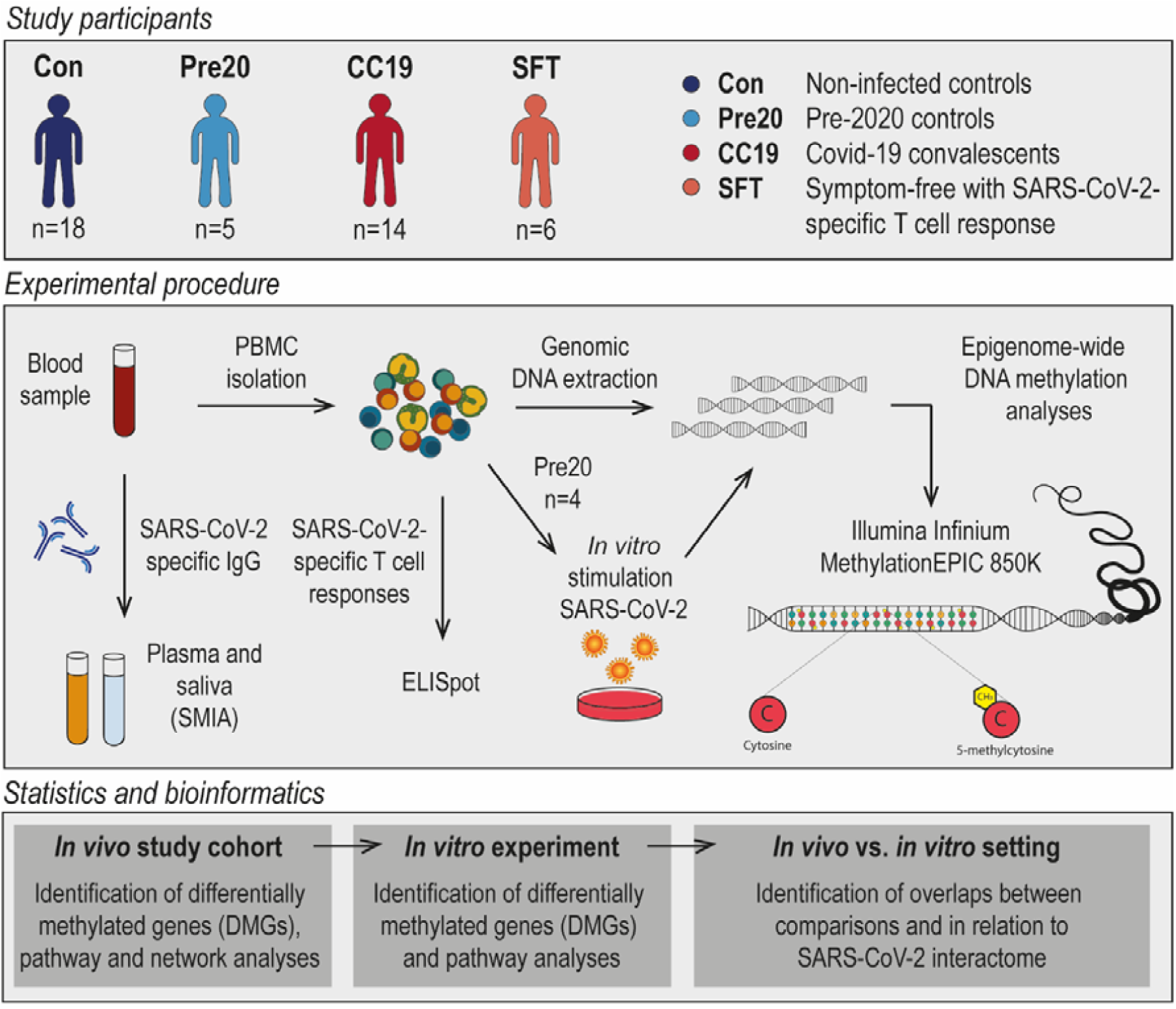
Outline of included participants, experimental procedures as well as statistical and bioinformatic approaches utilised in the present study. A brief overview of the present study. CC19 – convalescent COVID-19, Con – non-infected control, DMG – differentially methylated gene, Pre20 – Pre-2020 non-infected control, SFT – symptom-free individuals with SARS-CoV-2-specific T cell response, SMIA – suspension multiplex immunoassay.

### PBMC and plasma isolation from whole blood

Peripheral blood was collected in three 10 ml EDTA tubes (BD Vacutainer, 10331254, Fisher Scientific, Sweden). Up to 20 ml of whole blood was used for PBMC isolation after Ficoll-Paque Plus gradient centrifugation (GE17-1440-03, GE Healthcare Life Sciences, Sigma-Aldrich, Sweden) with SepMateTM tubes (85450, StemCell technologies, France) according to the manufacturer’s protocol. Cells were frozen in 10% DMSO (10103483, Fischer Scientific, Sweden) in fetal bovine serum (FBS) (10270106, Gibco, Fischer Scientific, Sweden) and kept at -150°C until analysis. After thawing, the cells were washed twice in cell culture medium (RPMI medium 1640, 31870-025, 10% fetal bovine serum, 1% penicillin/streptomycin, 15140, 1% L-glutamine, 25030081, all from Gibco, Fischer Scientific, Sweden) further on termed as complete culture medium, prior to further processing. Up to 10 ml of whole blood was used for plasma separation by centrifugation (2000g for 15min, 4°C) and aliquots were stored at -80°C till further analysis.

### Measurements of SARS-CoV-2-specific T cell responses using ELISpot

Peptides for the spike (S) protein of SARS-CoV-2 were obtained from Mabtech (3629-1, Sweden) and were reconstituted with di-methyl-sulphoxide (DMSO) at a concentration of 200 µg/ml according to the manufacturer’s instructions. The SARS-CoV-2 S1 scanning pool contains 166 peptides consisting of 15-mers, overlapping with 11 amino acids, covering the S1 domain of the spike S1 protein (amino acid 13-685). The peptides were combined into one pool. IFN-γ ELISpot Plus kit was purchased from Mabtech (3420-4HST-10, Sweden). Briefly, the pre-coated wells were plated with unfractionated PBMCs at counts of 300 000 cells/well, and the cells were cultured with peptides for the S protein of SARS-CoV-2 at a final concentration of 2 µg/ml (diluted in complete culture medium) for 20 to 22 hrs in a 37°C, 5% CO_2_ incubator. Cells cultured with medium alone were used as negative controls. Stimulation with anti-CD3 antibody at a concentration of 1 µg/ml was used as a positive control for each subject. Anti-CD28 antibody (3608-1-50, Mabtech, Sweden) was included at a final concentration of 0.1 μg/ml as a co-stimulator. All experiments were conducted in duplicates and results represent the mean of the duplicates. The plates were then processed according to the manufacturer’s protocol. Estimation of specific T cell numbers was expressed as spot-forming cells per 1×10^6^ PBMCs (SFC). SFC were counted using an automated reading system (BioSys Bioreader 5000 Pro-F beta, Bio-sys GmbH, Germany) and assessed with the Bioreader 5000 analyser. A stimulation index was calculated by dividing the SFC elicited by a SARS-CoV-2 stimulus by the SFC present in the negative control wells. An increment value was calculated by subtracting the SFC from the negative control wells from the SFC of the stimulated wells. A stimulus was considered to be positive when the stimulation index was >2, and the increment value was >10.

### Saliva samples

Prior to saliva collection, participants were required to rinse their mouth with water and confirmed they did not show documented oral disease or injury, that they had fasted, refrained from smoking, chewing a gum, taking oral medication, tooth brushing for a minimum of 1 hour before sampling and that no dental work had been performed within 24 hours prior to sample collection. Donors were asked to provide a 5 ml sample of saliva in a 50 ml sterile conical tube by passive drooling.

All saliva samples were stored/transported on ice upon receipt of the laboratory for processing to preserve sample integrity. Samples were centrifuged (2500g for 20 minutes at 4°C) to pellet cells and insoluble matter. The supernatant was collected and samples were complemented with cOmplete™ protease (#11836170001, Sigma) and PierceTM phosphatase inhibitor cocktails (#88667, Thermo Scientific), aliquoted and frozen/stored at -80°C on the same day. On the day of the assay, samples were thawed and micro-centrifuged (2500g for 10 minutes at 4°C) prior to analysis.

### Antibody responses in plasma and saliva using Suspension Multiplex Immunoassay (SMIA)

MagPlex-C microspheres (Luminex Corp., Austin, TX, USA) were used for the coupling of antigens according to the manufacturer’s protocol as previously described.^25^ Briefly, 200 µl of the stock microsphere solution (1.25 × 10^7^ beads/ml) were coupled by adding 10 μg of recombinant SARS-CoV-2 Spike protein RBD His-Tag (#40592-V08B, SinoBiological Inc., USA). After the coupling, beads were incubated in phosphate buffered saline (PBS: 0.15 M sodium chloride, 10 mM sodium phosphate, pH 7.4) containing 0.05% (v/v) Tween 20 (PBS-T) for 15 min on a rocking shaker at RT. The beads were then washed with 0.5 ml StabilGuard solution (SurModics, Eden Prairie, MN, USA, #SG01-1000) using a magnetic separator (Milliplex® MAG handheld magnetic separation block for 96-well plates, Millipore Corp. Missouri, USA. Cat. #40-285) and resuspended in 400 µl of StabilGuard solution. The coupled beads were stored at 4°C in the dark until further use.

For plasma samples, 50 µl of plasma diluted 1:1000, and for saliva samples 50 µl of sample diluted 1:2 in PBS-T containing and 1% (v/v) BSA (Sigma-Aldrich Sweden AB, Stockholm, Sweden, #Sigma-Aldrich-SRE0036) (PBS-T + 1% BSA) was added per well of a flat bottom, 96-well µClear non-binding microtiter plate (Greiner Bio-One GmbH, Frickenhausen, Germany, #Greiner-655906). Fifty microliters of a vortexed and sonicated antigen-coupled bead mixture suspended in PBS-T + 1% BSA (∼50 beads/µl) was then added to each well. The plate was incubated in the dark at 600 rpm for 1h at RT. The wells were then washed twice with 100 µl of PBS using a magnetic plate separator. The beads were resuspended in 100 µl of 1 µg/ml goat anti-human IgG-PE labelled antibody (Southern BioTech, Birmingham, AL, USA. Cat. #2040-09) in PBS-T + 1% BSA and incubated for 30 min at RT in the dark with rotation at 600 rpm. The beads were subsequently washed twice with PBS, resuspended in 100 µl of PBS and analysed in a FlexMap 3D® instrument (Luminex Corporation, Austin, TX, USA) according to the manufacturer’s instructions. A minimum of 100 events for each bead number was set to read and the median value was obtained for the analysis of the data. All sample analyses were repeated three times. A naked, non-antigen-coupled bead was included as a blank along with PBS-T + 1% BSA as a negative control.

### *In vitro* stimulation with SARS-CoV-2

PBMC samples from four healthy blood donors, frozen in 2019 in -150°C in foetal bovine serum (FBS) with 10% DMSO, were thawed and added to 10 ml of Gibco Dulbecco’s Modified Eagle Medium (DMEM) (Thermo Fisher Scientific, Waltham, US) containing 1% L-glutamine (Cat no: 25030-024, Gibco, Waltham, Massachusetts, USA), 1% penicillin-streptomycin (Cat no: 15140148 Gibco) and 10% normal human serum (NHS) (pooled from 5 donors) filtered through a 40 µm strainer and pre-heated to 37°C. The cells were washed two times by centrifugation at 330g for 10 min. The pellet was resuspended in 1.5 ml medium and 2 million per donor were seeded in six-well plates and incubated for 16-24 h. The cell culture media were collected, and centrifugated at 330g for 5 min to pellet the non-adherent cells.

For *in vitro* infection experiments, SARS-CoV-2 virus previously isolated in a Biosafety level 3 lab according to local safety regulations from the nasopharyngeal aspirate of a COVID-19 patient (early April 2020) was used.^26^ The isolated virus was passaged five times in Vero E6 cells and for cell infection experiments, freeze-thawed medium supernatants of 4-5 days infected cells or mock supernatants were used. Virus titers were determined using immunoperoxidase assay. In brief, two-day old confluent cells (in a 96-well plate) were first washed with Dulbecco’s Modified Eagle’s Medium (DMEM) (Gibco, Code: 13345364) containing 100 μg/ml gentamicin, and 100 μl of 10-fold serially diluted SARS-CoV-2 virus lysate was added in quadruplicate. SARS-CoV-2 or mock Vero cell supernatant was added to the PBMC cultures corresponding to a multiplicity of infection of 0.01. 2 hours post infection the cells were washed twice with DMEM and 100 μl of fresh DMEM (containing 2% FBS and 100 μg/ml gentamicin) was added, and the plate was incubated for 8 hours at 37^°^C in presence of 5% CO_2_. After incubation, the supernatant was discarded, and the cells were fixed for 2 hours with 4% formaldehyde. Next, Triton-X (1:500 in phosphate buffered saline, PBS) was added for 15 min, washed once with PBS and incubated for 2 hours at 37^°^C with PBS containing 3% BSA. Next, the cells were incubated with mouse-anti-dsRNA antibody (Scions, Code: J2 at 1:100 dilution) for 1.5 h followed by detection using horseradish peroxidase–conjugated goat anti-mouse IgG (heavy plus light chain) (Catalog: 1706516, Bio-Rad Laboratories, Hercules, CA, USA) (1:1000) for 1 h. The plates were washed five times with PBS between every incubation, all incubations were done at room temperature and the antibody dilutions were made in PBS containing 1% BSA. Finally, the SARS CoV-2 infected Vero E6 cells were identified using 3-aminoethylcarbazole (AEC) substrate. The spots representing virus-infected cells were counted under the light microscope and the virus lysate was titrated to be 5×10^6^ per ml.

Cells were monitored in the IncuCyte S3 live cell analysis system (Sartorious, Göttingen, Germany) to allow quantification of cell death in SARS-CoV-2 infected wells versus controls. After 48h incubation the cell culture media was collected from each well and centrifugated at 330g for 5 min to collect the non-adherent cells. Lysis buffer (RLT from the AllPrep® DNA/RNA Mini Kit, Qiagen, Hilden, Germany) was added to the wells to lyse adherent cells and the mixture was then added to the pelleted non-adherent cells in order to collect DNA (according to the manufacturer’s instructions) from the entire PBMC fraction.

### Epigenome-wide DNA methylation analyses

#### DNA extraction and quantification

For the performance of epigenome-wide DNA methylation analyses, DNA was extracted from the above isolated PBMCs (approximately 2×10^6^ cells) using the AllPrep® DNA/RNA Mini Kit (Cat no: 80204, Qiagen, Hilden, Germany) according to the manufacturer’s instructions. Concentrations of extracted DNA were measured using the Qubit® 4.0 Fluorometer (Thermo Fisher Scientific, Waltham, Massachusetts, U.S), using dsDNA High Sensitivity (HS) Assay Kit and RNA HS Assay Kit. The measurement was performed according to the manufacturer’s instructions.

#### Illumina MethylationEPIC 850K array

DNA samples were sent to the Bioinformatics and Expression analysis Core facility, Karolinska Institutet, Stockholm, Sweden, where the samples first went through bisulphite conversion on site, followed by the performance of the Illumina Infinium MethylationEPIC 850K array. 200 ng of DNA from each sample was analysed.

### Statistics

#### Descriptive analyses on demographic variables

Initial descriptive analyses of demographic variables were performed on the available information about age, gender, smoking and BMI (kg/m^2^). Continuous variables were compared using an unpaired two-tailed t-test and categorical variables were examined using the Pearson χ2 test or Fisher’s exact test (if the number of observations was smaller than five), see Table S1.

#### DNA methylation analyses

The resulting raw IDAT-files from the MethylationEPIC array analyses were processed in R programming environment (version 4.0.2). The analyses described below were identically performed for the clinical *in vivo* cohort and the *in vitro* experiment, unless stated otherwise.

#### Pre-processing and quality control

The resulting raw IDAT-files containing the raw DNA methylation profiles for each cell type were analysed in R (version 4.0.2) using the minfi package^27^ (version 1.36.0) and the data were pre-processed in several steps. The following filters were applied: i) removal of probes with detection p-values above 0.01, ii) removal of non-CpG probes, iii) removal of multi-hit probes, iv) removal of all probes in X and Y chromosomes.

##### In vivo

We removed the sex chromosomes from our data set, as female X-inactivation skews the distribution of beta values (Figure S1). Of the initial 865 918 probes, 841 524 probes remained upon filtering. After filtering, quality control was performed, and normalisation of the data was done with subset-quantile within array (SWAN) normalisation method.^28^ The β-values and M-values of the samples were calculated against each probe per sample. The quality of the data was assessed before and after the normalisation (Figure S2). Thereafter, we performed singular value decomposition (SVD) analyses using the ChAMP package^29^ (version 2.19.3) to identify underlying components of variation within the filtered and normalised data set (Figure S3). Significant components consisted of slide, batch and sample groups that contributed to variation within the data set. Corrections were performed for the identified components using ComBat from the SVA package^30^ (version 3.38.0). As PBMCs consist of multiple nucleated cell types in peripheral blood, we utilised the Houseman method to infer cell type proportions within the samples.^31^ No differences could be determined in cell type proportions between any of the individuals or between sample groups (Table S2), motivating our choice of not correcting for these cell type proportions.

##### In vitro

In this dataset, we did not have any information on demographic variables, as the samples derived from anonymous donors. However, we still removed the sex chromosomes from our data set, as female X-inactivation skews the distribution of beta values. Of the initial 861 728 probes, 837 694 probes remained upon filtering. After filtering, quality control was performed, and normalisation of the data was done with SWAN normalisation method.^28^ The Houseman method was utilised to infer cell type proportions within the samples^31^, yet again revealing no differences could be determined in cell type proportions between any of the individuals (Table S2), motivating our choice of not correcting for these cell type proportions. The β-values and M-values of the samples were calculated against each probe per sample. The quality of the data was assessed before and after the normalisation (Figure S4). SVA package (version 3.40) was applied to correct the batch effect. Cell deconvolution was performed using FlowSorted.Blood.EPIC package (version 1.11).

#### Differential DNA methylation analysis

##### In vivo

As we were interested in studying CpGs that were differentially methylated between CC19s and non-infected controls from both before and after the start of the COVID-19 pandemic, we performed differential DNA methylation analyses, using the limma package (version 3.46.0). A linear model was fitted to the filtered, normalised and SVD-corrected DNA methylation data. Identified sources of variation that were still present upon SVD correction provided the basis for the inclusion of these variables as co-variates in the models, in this case gender and BMI (Figure S3). For each investigated probe, moderated t-statistics, log2 Fold Change (logFC) and p-values were computed. The logFC values represent the average beta methylation difference (from hereon referred to as mean methylation difference, MMD) between the CC19s *vs*. non-infected controls (Cons + Pre20). Differentially methylated CpGs (DMCs) were defined as CpG sites having a nominal p-value of less than 0.01 along with an MMD of > 0.2. As a means to ascertain the quality of the identified DMCs, genomic inflation and pertaining bias were estimated using the BACON package^32^ (version 1.18.0). As the estimated genomic inflation for the comparison was close to 1 (genomic inflation: 1.20, bias: 0.01, Figure S5), this suggested that no major genomic inflation was present in the comparisons, and no correction for this was deemed necessary. The distribution of the DMCs among all investigated DNA methylation sites were illustrated by creating volcano plots (EnhancedVolcano, version 1.8.0). Thereafter, the DMCs were mapped to their corresponding DMGs. DMGs contained at least one DMC, and were considered hyper- or hypomethylated if all DMCs within the gene were hyper- or hypomethylated, respectively. If both hyper- and hypomethylated genes were present in the same gene, the gene was considered having a mixed methylation pattern.

##### In vitro

To evaluate the difference between the mock and infected sampeles, the fold change was calculated using the cut-off obtained from the density plot (M-value >|2|; Figure S6) for each CpG site. Only those CpGs with higher values than the cut-off, were selected for further analysis. Venn analysis was performed among the samples using the ggVennDiagram (version 1.1) package in R (version 4.0.3) and bioconductor (version 3.12).

#### Pathway over-representation analyses

To make biological sense of the putatively SARS-CoV-2-induced DNA methylation differences, we performed PANTHER pathway over-representation test analyses using the PANTHER database (version 16.0). The Fisher’s exact test was used for generation of nominal p-values (significance level set to p-value of < 0.05), in case false discovery rate correction was too stringent. The significantly enriched pathways were displayed in dot plots generated in R using ggplot2 package (version 3.3.3).

#### Network analyses

Network analyses were conducted to generate further and wider biological insight about the DMGs generated in the *in vivo*. An input object was constructed using the pre-2020 (Pre20, n=5) and post-2020 (Con, n=18) non-exposed controls and COVID-19 convalescents (CC19, n=14), as a two-column data frame containing gene annotation and P-value of the significant DMGs (n=54). The graph clustering algorithm MCODE^33^ was used to identify molecular complexes and create a large disease module, which was then fitted to a protein-protein interaction network, and both were analysed and rendered in Cytoscape (version 3.8.0). High confidence interactions with a STRINGdb confidence value >0.7 were displayed in the network. Centrality measurements of degree, betweenness and closeness were used to expose the most central nodes in the network. Finally, a functional enrichment of the genes present within the module was carried out using StringDB.^34^ In addition, the inference of modules was performed with two other methods from the MODifieR package (DIAMOnD and WGCNA)^35^ to study whether it was possible to condense the module genes to fewer genes of particular interest within the network, for both the *in vivo* and the *in vitro* setting.

#### Overlap to SARS-CoV-2 interactome

A publicly available protein-protein interaction (PPI) network of SARS-COV-2 and human genes curated by BioGRID (version 4.4.197) was downloaded from the Network Data Exchange in Cytoscape (version 3.8.0). The DMGs from the *in vivo* and *in vitro* setting alongside the gene list from the module generated by MCODE were overlapped onto the PPI network to visualise their respective distributions.

## RESULTS

### COVID-19 convalescents display altered DNAm patterns compared to non-infected controls

We compared epigenome-wide DNAm patterning in peripheral blood mononuclear cells (PBMC) from non-infected controls (Con, n=19), COVID-19 convalescents who had recovered from mild or moderate symptoms (CC19, n=14), donor blood collected before the pandemic (Pre20, n=5) and from asymptomatic individuals presenting with SARS-CoV-2-specific T cell responses (SFT, n=6, Figure 1). Comparisons of demographic variables revealed no significant differences between any of the groups (Table S1). To examine any inherent differences in the DNA methylome between the different sample groups, principal component analyses (PCA) were performed. Three principal components (PC) were identified as both contributing to the variation within the DNAm data and correlating with the sample groups (Figure S7). A three-dimensional illustration of these three most contributing components revealed that the CC19 subjects are distinct from the Con, Pre20 and SFT subjects, whose centroids clustered more closely together (Figure 2a, Figure S8). The observed methylome-wide differences prompted us to identify differentially methylated CpGs (DMCs), which we defined as CpG sites with a nominal p-value of <0.01 along with a mean methylation difference (MMD) of >0.2. We found 87 DMCs, when comparing the DNA methylomes of CC19s to the merged groups of Cons and Pre20s (Figure 2b, Table S3a). This identified DMC signature could furthermore distinguish the CC19s from Cons, Pre20s and SFTs (Figure 2c), suggesting that a past SARS-CoV-2 infection may have resulted in modulation of the epigenome that persists at least a couple of months after the virus is eliminated from the body.

**Figure 2.**
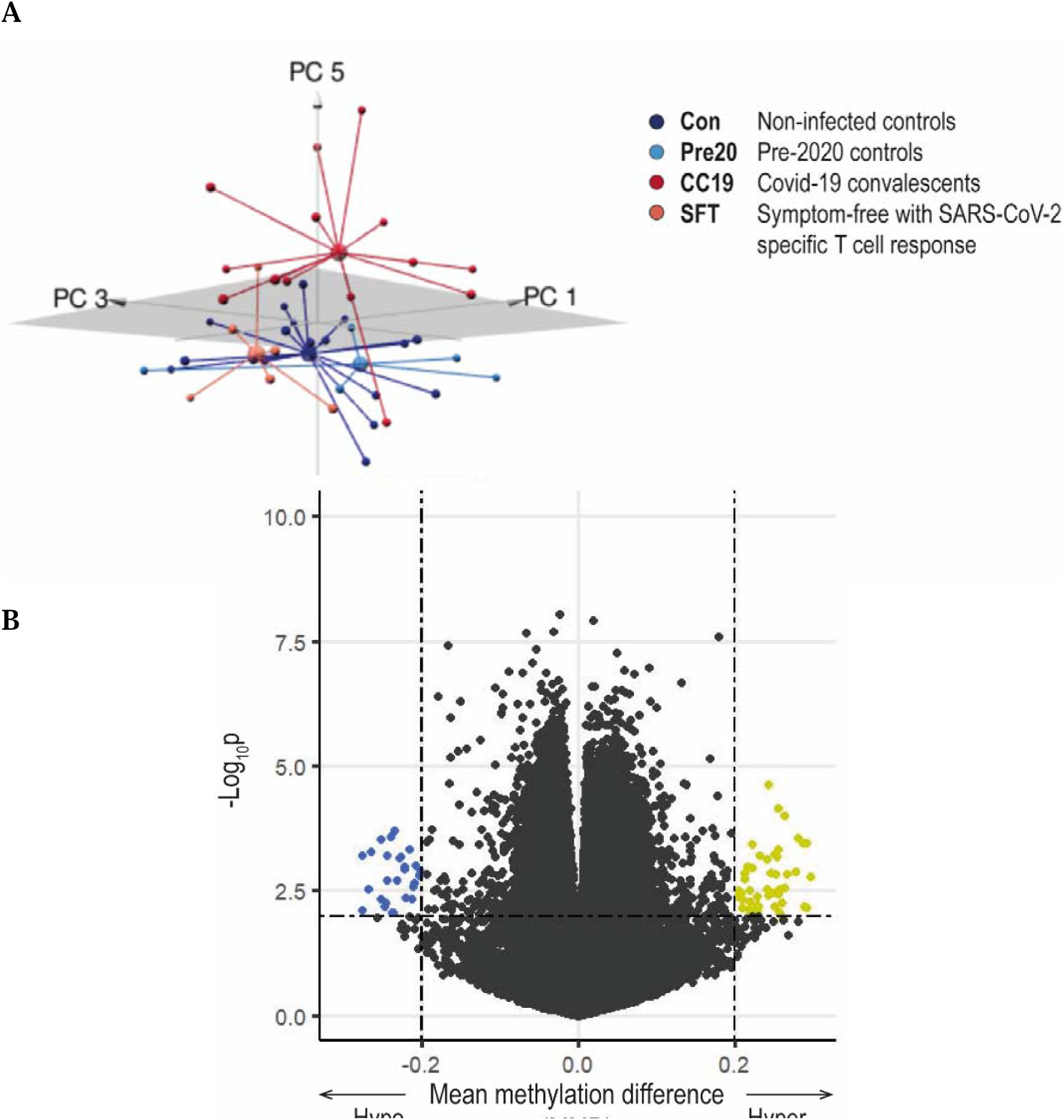
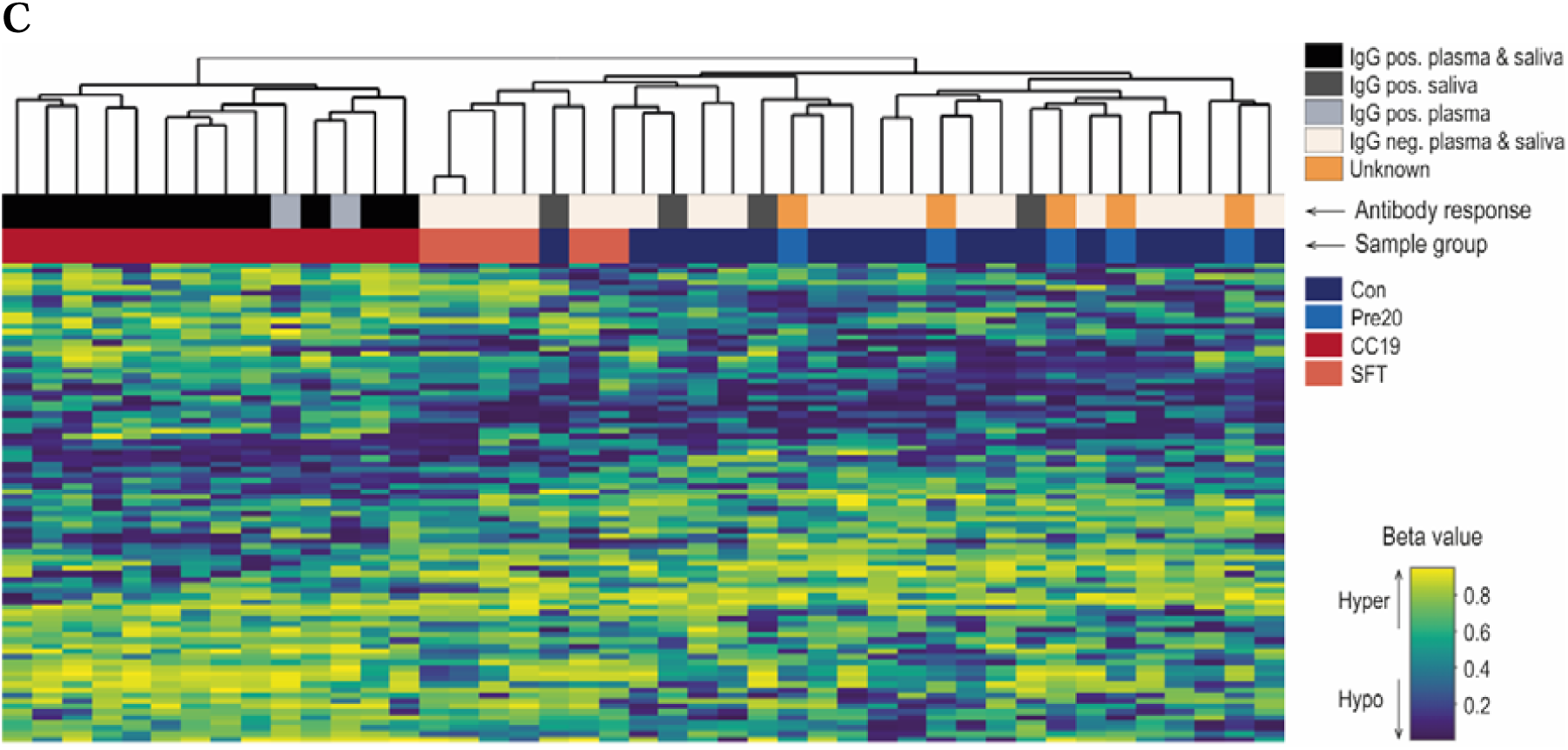
Principal component and differential DNAm analysis of PBMC DNA methylomes in COVID-19 convalescents and uninfected controls. Upon filtering and normalisation, the DNAm data were subjected to PCA. Panel A shows a 3D-PCA plot of principal components (PC)1, PC3 and PC5, where the group means are illustrated as centroids. DMCs were identified comparing CC19s to Cons and Pre20s by computing a linear model on the DNAm data. Panel B illustrates a volcano plot of the CC19 *vs*. Con + Pre20 DNAm data. The dash-dotted horizontal line represents a nominal p-value cut-off of 0.01, and the vertical lines represent a cut-off in mean methylation difference (MMD) in CC19 *vs*. Con + Pre20 of > ± 0.2. Panel C shows a heatmap representing an unsupervised hierarchical clustering analysis of individual β values of the 87 identified DMCs in B. The individuals’ antibody status is indicated as a grey-scale (unknown = anonymous Pre20 blood donors, orange).

Interestingly, a majority of CC19s showed positive SARS-CoV-2-specific IgG responses both in the circulation and in saliva (Figure 2c). Individuals who showed T cell responses towards SARS-CoV-2 or presented with SARS-CoV-2-specific antibodies in saliva while being negative for antibodies in plasma, aligned with the uninfected controls in the PCA and unsupervised clustering analyses (Figure 2b-c).

### Differential methylation of COVID-19 convalescents identifies a putatively SARS-CoV-2-induced module

To further explore the biological impact of SARS-CoV-2 exposure in the CC19 subjects, the identified DMCs were annotated to their respective differentially methylated genes (DMG), resulting in 54 unique genes (Table S3b). Subsequent pathway over-representation analyses revealed involvement in two significantly over-represented pathways (Wnt and integrin signalling pathways, Table S4).

As a means to elaborate on the wider interaction context in which the DMGs act with other proteins, the DMGs (n=54) were used as seed genes in the identification of SARS-CoV-2-induced modules in network analyses. The resulting module consisted of 66 genes from the protein-protein interaction network, with 139 interactions, which is significantly more interactions than expected (34 interactions) for a network of that size (Figure 3, Table S5). Six of these genes were present in at least two module identification methods (*INS, HSPA4, SP1, ESR1, TP53* and *FAS*), and they were all located in the centre of the module. The four genes with the highest combined centrality scores were *HSP90AA1, TP53, INS* and *CFTR*. Pathway over-representation analyses of the 66 module genes revealed involvement in pathways such as apoptosis signalling, muscarinic acetylcholine receptor 1 and 3 signalling and gonadotropin-releasing hormone receptor pathway (Figure S9).

**Figure 3.**
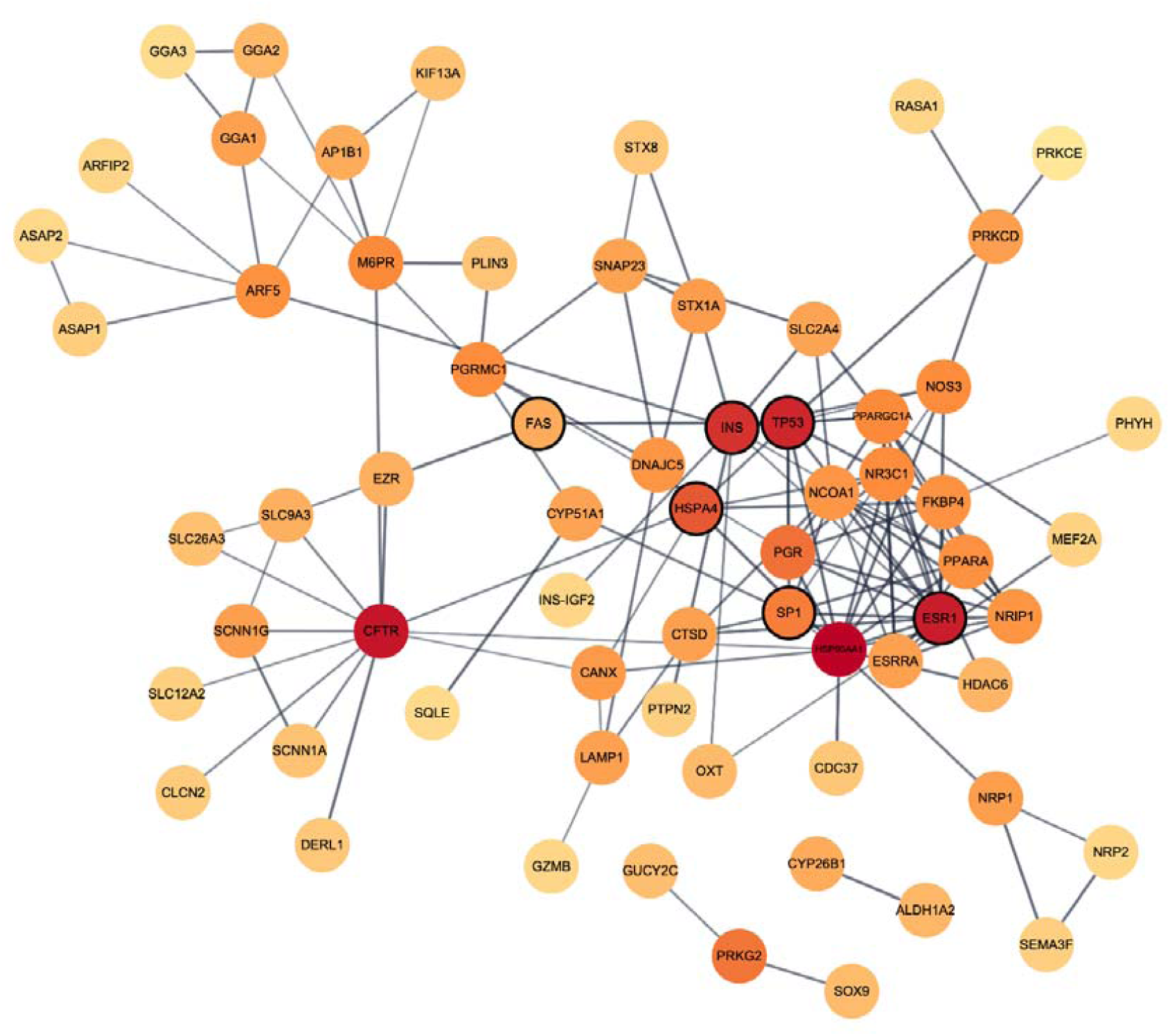
Network illustration of SARS-CoV-2-induced module genes from the *in vivo* comparison. A network module constructed by means of the graph clustering algorithm MCODE with the 54 DMGs from the *in vivo* setting as input. Nodes (n=66) represent genes and connecting lines represent high-confidence protein-protein interactions within the network (STRING combined score > 0.7). Combined ranked scores of centrality quantification of degree, betweenness and closeness is visualised as a colour (light orange to dark red) continuum, with dark red nodes constituting the most central parts of the network. Nodes that were also found both when utilising two other module identifying methods (DIAMOnD and WGCNA) and when performing the same analyses on the *in vitro* data set using MCODE are enclosed with a black line.

### SARS-CoV-2 stimulated PBMCs *in vitro* reveal overlaps with *in vivo* differential methylation, network analyses and SARS-CoV-2 interactome

In the present study, we only had access to self-reported time-after-onset of COVID-19 symptoms (Table S6), thus making the immediate effects of SARS-CoV-2 exposure on the epigenome impossible to analyse. Moreover, as the virus-induced DNAm patterns in the CC19’s may fade over time, we set out to examine SARS-CoV-2-induced DNAm patterns in an *in vitro* setting. To this end, we exposed PBMCs collected from blood donors in 2019 to SARS-CoV-2 at a low multiplicity of infection for 48h to mimic immediate *in vivo* exposure to the virus (Figure S10). Exploring the intra-individual DNAm differences between stimulated and unstimulated cells, a set of DMCs (n=3693, Figure 4) were identified to be shared between all four individuals (Table S7a-b). These DMCs mapped to in total 606 DMGs (542 unique genes, Table S7c), which were significantly over-represented in a number of pathways including several glutamate receptor pathways, muscarinic acetylcholine receptor 1 and 3 signalling pathway, as well as the Wnt and cadherin signalling pathways (Figure S11).

**Figure 4.**
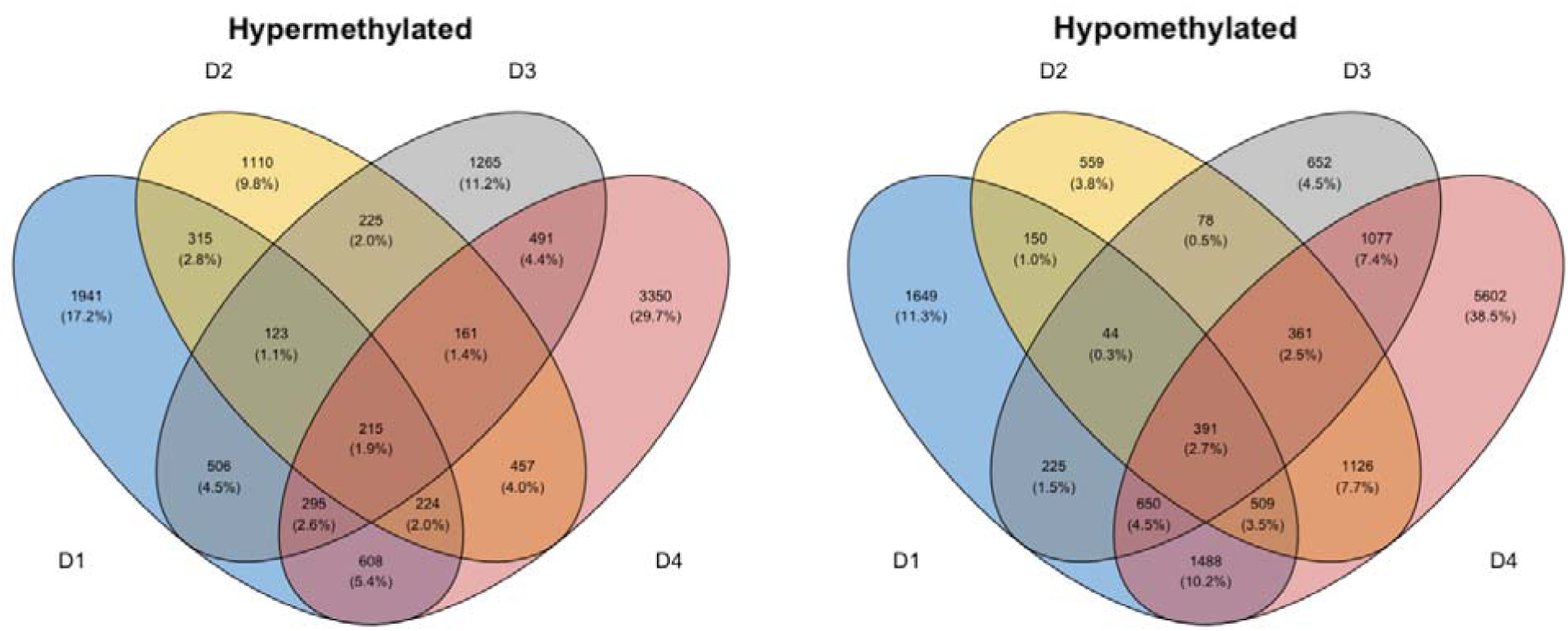
Differential DNAm analyses of PBMCs stimulated *in vitro* with SARS-CoV-2. Venn diagrams depicting the overlap of DMCs from the SARS-CoV-2 *in vitro* stimulated PBMCs. Intra-individual comparisons of differential DNAm were performed in treated *vs*. untreated PBMCs from four different blood donors (D1-D4) collected before the start of the COVID-19 pandemic (2014-2019). DMCs were defined as a fold change in M-value >|2|. These DMCs were further mapped to their corresponding annotated genes (DMGs, n=542).

As similar pathways were revealed in the findings from the *in vivo* study and the SARS-CoV-2 stimulations, we wanted to explore further similarities in DNAm between the *in vivo* and *in vitro* settings. Analyses of the overlap of shared DMGs identified in the two comparisons revealed eight overlapping DMGs (*OR12D3, PCSK6, INPP5A, RAD51B, CDH4, PHACTR3, CDH13, SFTA2*). Additionally, to understand the biological context of the genes identified in the *in vitro* comparison, we performed network analyses in the same manner as for the *in vivo* comparison. These analyses found a module consisting of six genes (*TP53, INS, HSPA4, SP1, ESR1* and *FAS*), which were among the previously identified module genes from the *in vivo* setting and were also identical to those that had been identified by more than two module identification methods (Figure 3). Furthermore, explorations of the overlap between identified genes in the differential DNAm analyses and network module analyses to the genes from a publicly available SARS-CoV-2 interactome identified numerous interactions in the *in vivo* (n=11/54), *in vitro* (n=100/542) and network module setting (n=33/66) (Figure S12, Table S8).

## DISCUSSION

The epigenetic events triggered during a mild COVID-19 disease course are largely unexplored, despite the fact that these individuals make up a majority of all SARS-CoV-2-infected individuals. The main finding of our study was an observed DNAm signature that was evident several months after recovery in CC19s compared to non-infected individuals. Although this has not, to our knowledge, previously been described, further investigations need to prove whether this particular signature is a remnant from the time of active infection. Studies of DNA methylomes in circulating cells of COVID-19 patients have so far focused on the hospitalisation phase for moderate-to-severe disease or at discharge^36-40^, and none of these studies report comparisons upon convalescence from a mild-to-moderate disease course. Not surprisingly, most of these studies mainly identify engagement of several antiviral immune-related pathways as well as inflammatory responses in severely ill COVID-19 patients compared to controls.^39, 40^ In contrast, our pathway over-representation analyses revealed the involvement of distinct, previously unappreciated pathways such as the Wnt signalling and the muscarinic acetylcholine receptor 1 and 3 signalling pathways. The Wnt signalling pathway has been implicated in several aspects of COVID-19, including development of inflammation, cytokine storms, as well as pulmonary fibrosis.^41^ Furthermore, potential viral hijacking of host Wnt targets has been suggested upon SARS-CoV-2 infection in multi-omics studies.^42^ The muscarinic acetylcholine receptor 1 and 3 signalling pathway was present in the module identification analyses from both the natural *in vivo* exposure and the *in vitro* stimulations. Interestingly, in post-viral fatigue patients, including post-SARS-CoV and myalgic encephalomyelitis/chronic fatigue syndrome patients, this signalling pathway is dysfunctional, which has been tentatively attributed to the development of anti-muscarinic receptor autoantibodies.^43, 44^ Although this was not investigated in our study, this could suggest that these pathways found may be implicated in the development of post-acute COVID-19 syndrome, as the effects we observe have persisted for months after the initial exposure to the virus.

In the present study, DNA methylome analysis of PBMCs identified a number of genes that were shared between the natural *in vivo* infection and following *in vitro* stimulation, which were further confirmed by several module identification methods. One of these genes was tumour protein 53 (TP53), an evolutionarily conserved protein that is one of the most well-studied hub genes in cell signalling due to its central role in cancer.^45^ TP53 has in several other studies previously been identified been identified as a hub gene, in whole blood from COVID-19 patients^46^, and been shown to interact with ACE2 in SARS-CoV-2-infected human induced pluripotent stem cell-derived cardiomyocytes.^47^ Moreover, transcriptomic analyses of PBMCs from a small group of patients infected with SARS-CoV-2 revealed involvement of apoptosis and p53 signalling pathways^48^, a finding that was further supported by studies of the SARS-CoV-2 interactome, where TP53 was identified as a central player in apoptosis-mediated pathways.^49^ Two additional genes, both members of the heat shock protein family, *HSP90AA1* and *HSPA4* stand out in the network derived from our *in vivo* and *in vitro* data. Interestingly, reports on differentially expressed genes overlapping between acute respiratory distress syndrome and venous thromboembolism datasets identified *TP53* and *HSP90AA1* as central genes, among the top ranked hub genes in their networks.^50^ *HSP90AA1* was previously shown to be upregulated in bronchial cells of patients with mild COVID-19 disease, as compared to those with a severe disease course^51^, suggesting that this gene may be of particular importance in the mounting of a protective antiviral response. Although our study does not provide any evidence for a protective role of the observed epigenetic alterations, HSP70 family members have been discussed as both anti-viral defence components^52, 53^, as well as anti-viral drug targets, against SARS-CoV-2.^54^ Altogether, our findings on the network centrality of the hub genes that we derived from the *in vivo* and *in vitro* data suggests that they may be of particular importance in the interaction with epigenetically modulated genes upon SARS-CoV-2 infection. Nevertheless, further studies are needed to elucidate the mechanistic role of these genes during infection and recovery from COVID-19.

A limitation of the study is the lack of validation of the DNAm findings on a transcriptional level. Hence, whether the observed DNAm patterns are indeed associated or causally linked to host protective or host detrimental immune responses still needs to be addressed in future studies, with more well-designed, larger cohorts and consecutive sample materials from the onset of SARS-CoV-2 infection. The investigation of epigenetic modifications in mild-to-moderately ill COVID-19 patients enabled us to discern DNAm differences that otherwise would have been masked by overriding inflammatory responses. Though these subtle changes may not primarily be relevant to immune response severity towards SARS-CoV-2, they may be insightful for the identification of both effective host protective mechanisms at play, or ensuing deliberating conditions such as long-COVID. The presentation of longstanding symptoms in long-COVID could be attributed to detrimental alterations in DNAm patterns, though originally triggered as a short-term anti-viral response.

In conclusion, we found epigenome-wide differences in DNAm patterns of individuals that had recovered from a mild-to-moderate disease course of COVID-19 compared to non-infected controls. This study suggests that DNAm is one of several epigenetic mechanisms that is altered upon SARS-CoV-2 infection, and further investigations should elaborate on whether they are induced by protective host responses or constitute virally-induced hi-jacking processes. Pinpointing these matters will aid the development of efficacious diagnostic tools and treatments of COVID-19 in the future.

## Supporting information

incl. Figures S1-S12, Tables S1 and S4-S6. Tables S2-S3 and S7-S8 are supplied in separate files.

## Data Availability

The datasets used and/or analysed in the presented work will be available upon publication on GeneExpression Omnibus, due to a pending patent application. The datasets comprise filtered and preprocessed DNA methylation data from deidentified individual samples in the study. The dataset will until publication be available using a secure token, provided by the authors upon request. Please, refer to GEO-ID: GSE178962 for further information on the data set.
Utilised scripts for performing the described statistical analyses within the paper, as well as for creating graphs, will be available on the following GitHub account upon publication (https://github.com/Lerm-Lab/Covid19).

## ACKNOWLEDGMENTS

We would like to thank the Bioinformatics and Expression analysis Core facility at Karolinska Institutet for their fruitful collaboration and the performance of the Illumina Infinium MethylationEPIC 850K arrays described in this paper. Additionally, we acknowledge the Swedish National Infrastructure for Computing (SNIC) at National Supercomputing Centre (NSC), Linköping University for the computing systems enabling the data handling, partially funded by the Swedish Research Council (Grant No. 2018-05973).

## AVAILABILITY OF DATA AND MATERIALS

The datasets used and/or analysed in the presented work will be available upon publication on GeneExpression Omnibus, due to a pending patent application. The datasets comprise filtered and preprocessed DNA methylation data from deidentified individual samples in the study. The dataset will until publication be available using a secure token, provided by the authors upon request. Please, refer to GEO-ID: GSE178962 for further information on the data set. Utilised scripts for performing the described statistical analyses within the paper, as well as for creating graphs, will be available on the following GitHub account upon publication (https://github.com/Lerm-Lab/Covid19).

## AUTHOR CONTRIBUTIONS

*Conceptualisation:* M.L. – equal, A.R. - equal, *Data curation:* S.Sa. – lead, J.D. – support, J.H. – support, *Formal analysis:* S.Sa. – lead, J.D. – support, L.K. – support, L.P. – support, J.H. – support, *Funding acquisition:* M.L. – equal, A.R. – equal, *Investigation:* E.A. – equal, M.R. – equal (ELISpot/SMIA), M.L. – equal, L.K. – equal, S.Sh. – support (*in vitro* SARS-CoV-2 stimulation of PBMCs), L.K. – lead (DNA extraction PBMC samples), *Methodology:* M.L. – equal, L.K. – equal, A.R. – equal, E.A. – equal, M.R. – equal, *Project administration:* M.L. - equal, A.R. – equal, J.H. – equal, S. Sa. – equal, *Resources:* A.R. – equal, M.L. - equal, *Software:* S.Sa. - lead, J.D. – support, L.K. – support, L.P. – support, *Supervision:* M.L. - equal, A.R. – equal, J.H. – equal, S. Sa. – equal, *Validation:* Not applicable, *Visualisation:* S.Sa. – equal, J.H. - equal, L.K. – support, J.D. – support, L.P. – support, *Writing – original draft:* J.H. – lead, all other authors - support, *Writing – review and editing:* J.H. – lead, all other authors – support.

