## Supplementary material for "Epigenome-wide DNA methylation profiling of healthy COVID-19 recoverees reveals a unique signature in circulating immune cells": incl. Figures S1-S12, Tables S1 and S4-S6. Tables S2-S3 and S7-S8 are supplied in separate files.

*Shared first authorship

**Corresponding author

^1^Division of Inflammation and Infection, Department of Biomedical and Clinical Sciences, Linköping University, Linköping, Sweden

^2^Division of Cell Biology, Department of Biomedical and Clinical Sciences, Linköping University, Linköping, Sweden

^3^Division of Molecular Medicine and Virology, Department of Biomedical and Clinical Sciences, Linköping University, Sweden

**SUPPLEMENTARY INFORMATION**

**incl. Figures S1-S12, Tables S1 and S4-S6. Tables S2-S3 and S7-S8 are supplied in separate files.**

**FIGURES**

**Figure S1. Density plot of beta value distribution between males and females.**
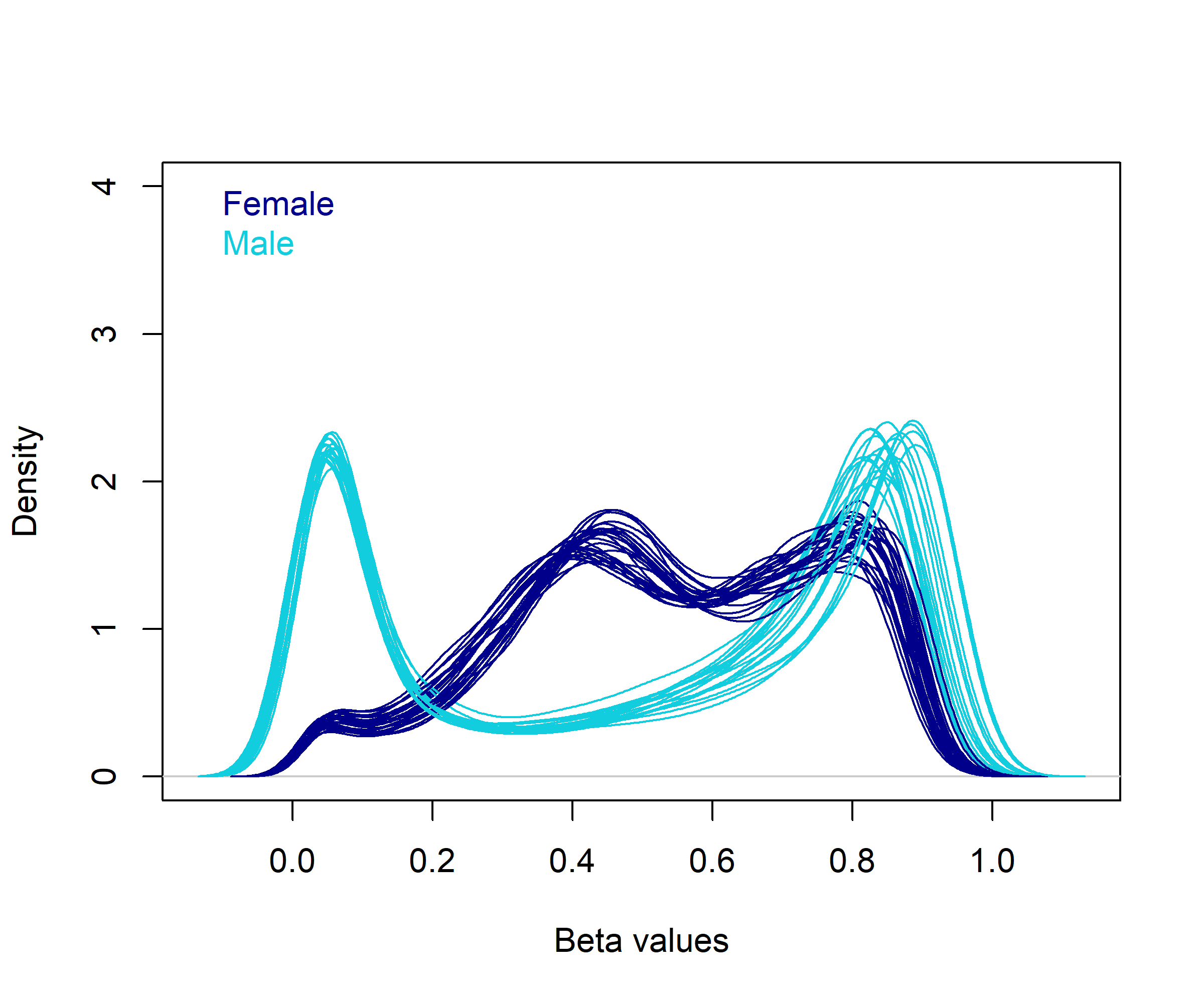

**Figure S1.** Graphical representation of the density of the beta value distribution on the X chromosomes of males and females in raw DNA methylation data. Female samples are illustrated in dark blue, males in light blue.

**Figure S2. Density plot of beta value distribution before and after normalisation.**
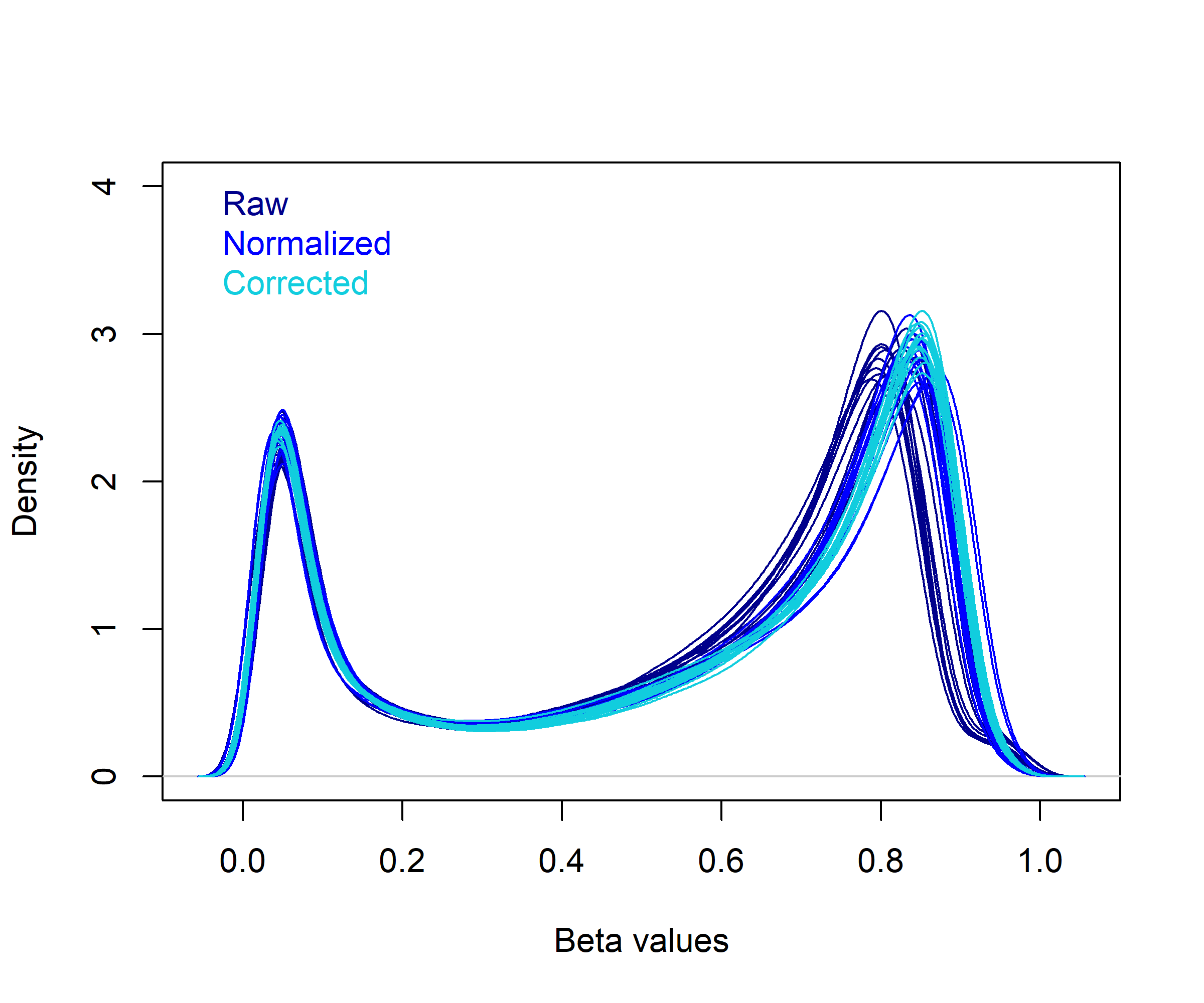
**Figure S2.** Graphical representation of the density of the beta value distribution on the autosomal chromosomes prior to and after normalisation and correction. Raw DNA methylation data are depicted in dark blue, SWAN-normalised data in lighter blue and batch corrected data are illustrated in coral blue.

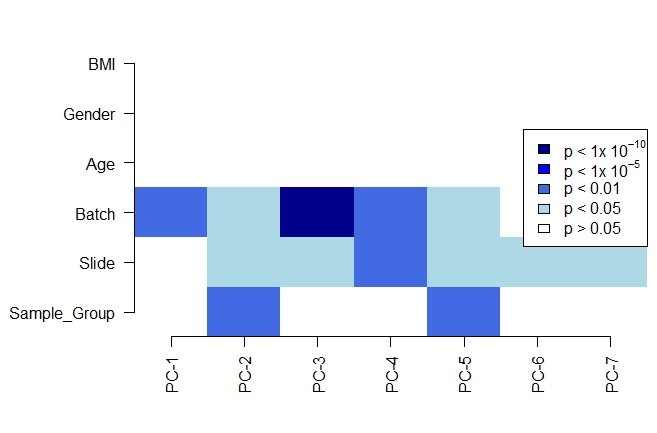

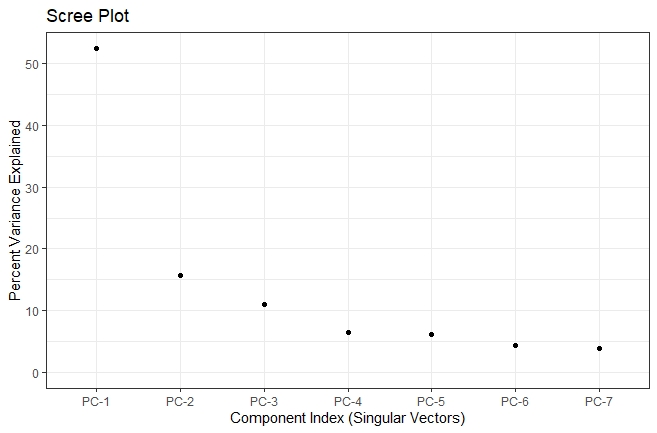

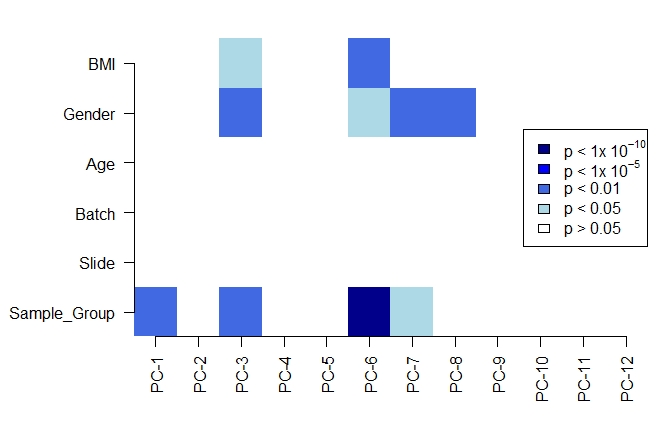

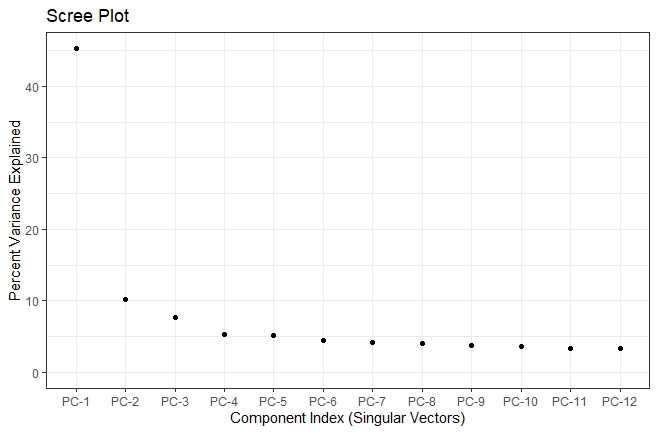
**Figure S3. Graphical representation of SVD analyses and scree plots before and after correction of batch effects.**

**a	Before correction**

**b	After correction**

**Figure S3.** Graphical representation of Singular value decomposition (SVD) analyses for identification of major components of variation using the ChAMP package. Analyses on the filtered and SWAN normalised data are displayed in panel a, and panel b shows the data upon correction for batch effects (batch and slide). The bottom panels present the % variance explained by each of the illustrated components in the SVD analyses.

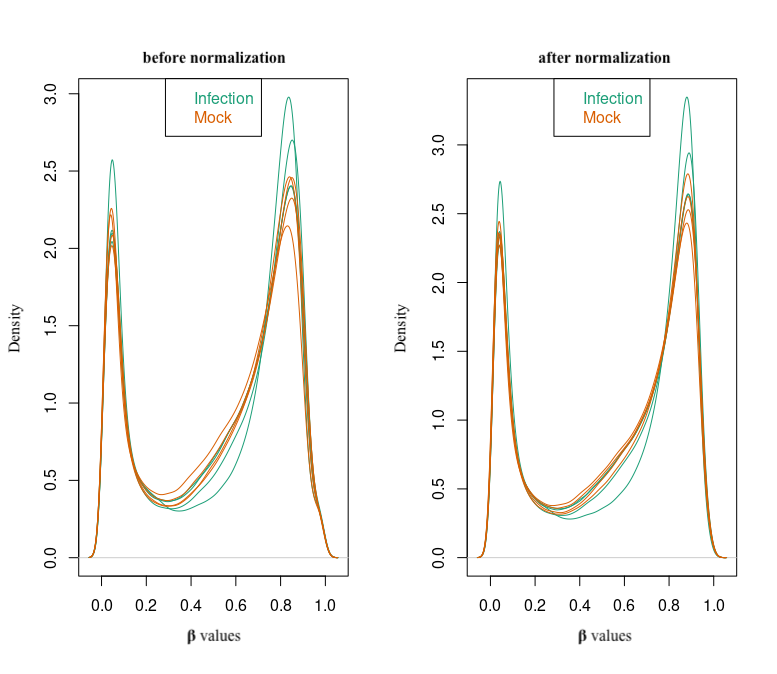
**Figure S4. Density plot beta distribution *in vitro* before and after normalisation.**

**Figure S4.** Graphical representation of the density of the beta value distribution on the autosomal chromosomes prior to (left panel) and after SWAN normalisation (right panel) using the SVA package. SARS-CoV-2 infected samples are illustrated in green, and corresponding non-infected mock samples in green.

**Figure S5. QQ-plot estimating genomic inflation and pertaining bias of the *in vivo* differential DNA methylation analyses.**

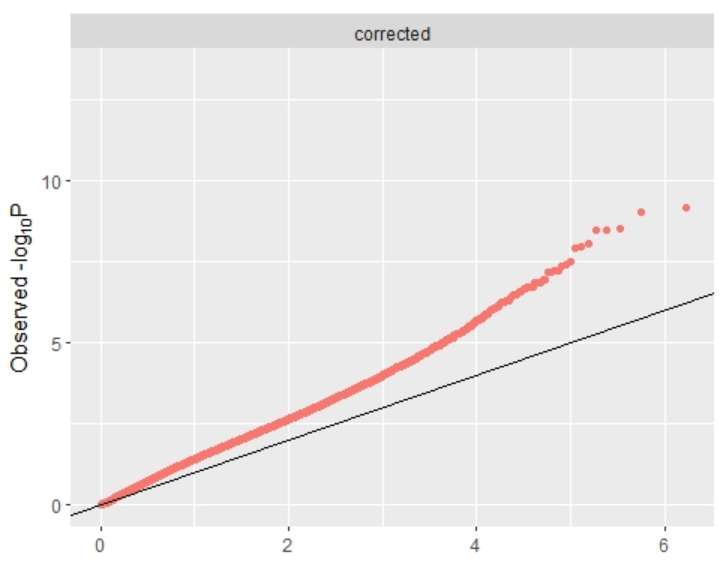

**Figure S5.** A QQ-plot illustrating the observed *vs.* expected –log10 of the obtained p-values from the differential DNA methylation comparison between CC19 and Con+Pre20 samples. The accompanying estimated values of genomic inflation and bias were 1·2 and 0·01, respectively. Calculations were performed using the R package BACON.

**Figure S6. Density plot of M-values to determine the cut-off for acquiring DMGs in the *in vitro* study.**

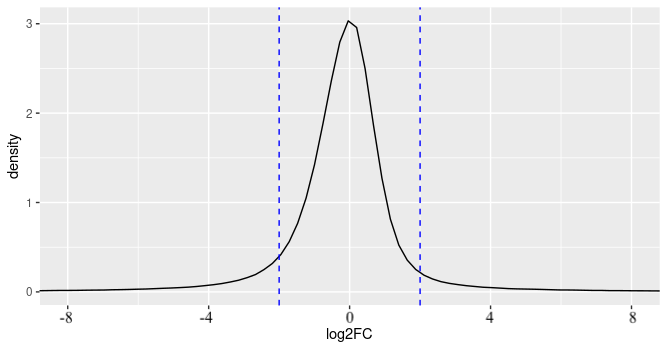
**Figure S6.** A density plot of the M-values, which was used to determine the cut-off for acquiring differentially methylated genes from the *in vitro* PBMC stimulation with SARS-CoV-2.

**Figure S7. PCA analysis of DNA methylomes**

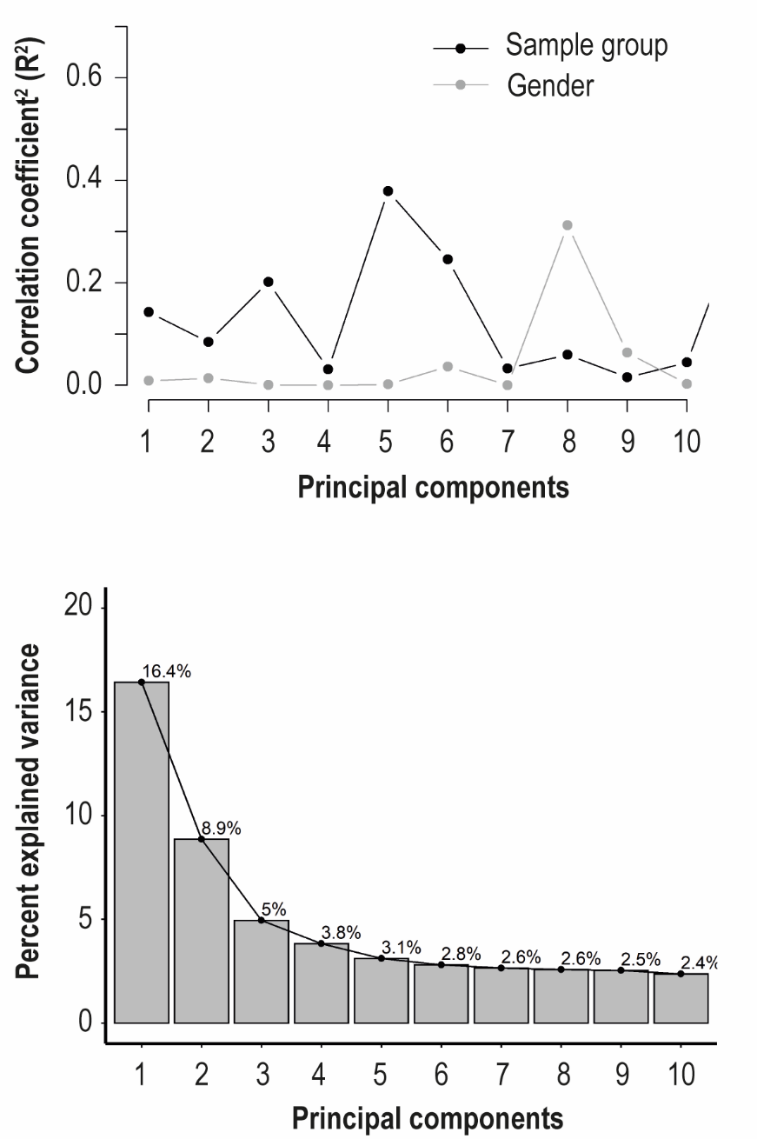
**a**

**b**

**Figure S7.** Upon filtering and normalisation, the DNAm data were subjected to PCA analysis. Panel a shows a correlation plot of the PCA-derived eigenvalues and the DNAm group data projected as Con/Pre20/CC19/SFT and male/female. In b. a scree-plot shows degree to which the identified components contribute to the variation observed within the DNAm data.

**Figure S8. A 3D-PCA animation illustrating three components that contribute to and correlate significantly with the sample groups in the *in vivo* data set.**

<https://drive.google.com/file/d/1OaBE73aQKm4XyxgtAA4Glo6BpE5q63B7/view?usp=sharing>

**Figure S8**. The PCA was performed on epigenome-wide DNA methylome data from peripheral blood mononuclear cells (PBMCs) upon filtering, normalisation and correction for underlying variation comparing non-infected controls (Con), convalescent Covid-19 (CC19) individuals, pre-2020 non-infected controls (Pre20) and individuals revealing a SARS-CoV-2-specific T cell response, despite reporting no symptoms (SFT). Follow link for a view of the .gif-file.

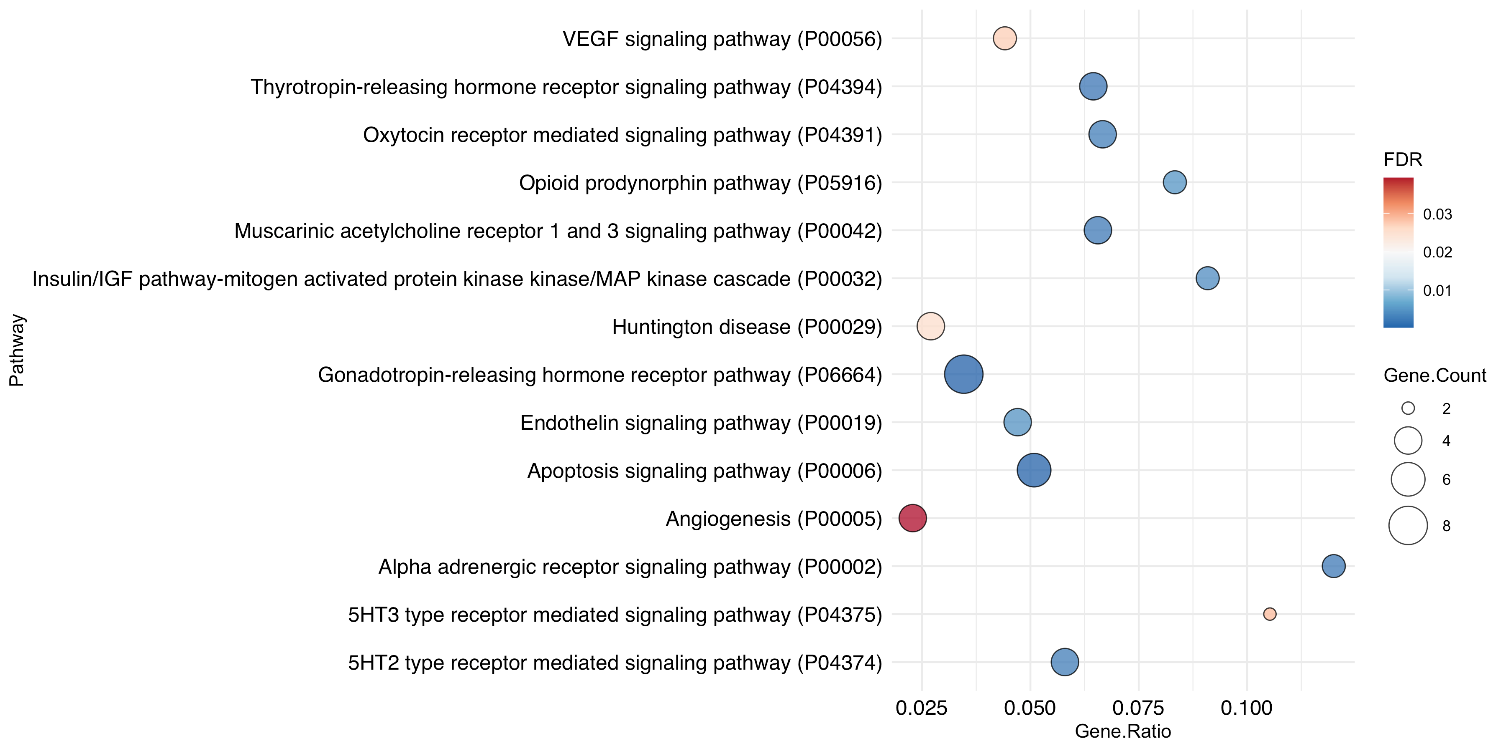
**Figure S9. Pathway over-representation analysis of network genes from the *in vivo* setting**

**Figure S9.** Results from pathway over-representation analyses of the 66 identified network genes in the protein-protein interaction network using PANTHER. Pathways with an FDR-corrected p-value < 0.05 were considered significant.

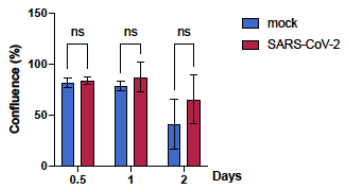
**Figure S10. Degree of confluence from SARS-CoV-2 *in vitro* stimulated PBMCs**

**Figure S10.** PBMCs from pre-2020 non-infected individuals (n=4) were stimulated with SARS-CoV-2 *in vitro* for 48h with a multiplicity of infection of 0.01. Differences between SARS-CoV-2 infected (red) and non-infected mock samples (blue) from the same individuals were compared using a two-way ANOVA.

**Figure S11. Pathway over-representation analysis of DMGs from the *in vitro* setting**
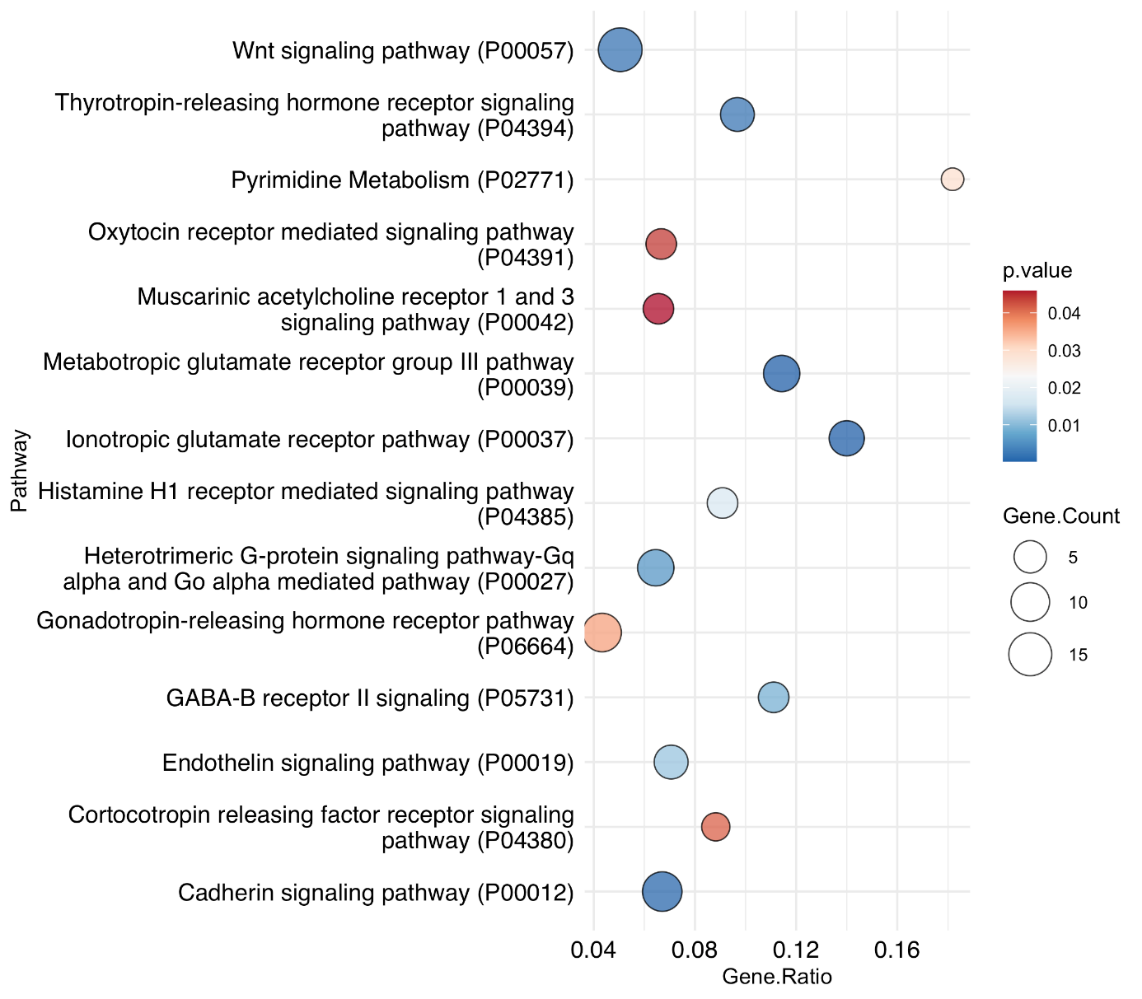

**Figure S11.** Results from pathway over-representation analyses in PANTHER based on the 542 DMGs originating from the SARS-CoV-2 *in vitro* stimulated PBMCs compared to non-stimulated PBMCs are shown. Pathways with a nominal p-value < 0.05 were considered significant.

**Figure S12. Overlap of genes from DNAm and network analyses to the SARS-COV-2 interactome.**

<http://www.ndexbio.org/#/network/5ac7c6bb-dac0-11eb-b666-0ac135e8bacf?accesskey=851c70f8967f4cbe6e02e992a9372c9ac4fe6a34a38b5d0a828b82efc93ee0a9>

**Figure S12.** Differentially methylated genes deriving from the DNAm analyses of the *in vivo* and *in vitro* setting as well as genes from the protein-protein interaction network analyses were compared to the large between-species interaction network consisting of all of the human genes that are known to interact with the different SARS-COV-2 proteins (as curated by BioGRID). SARS-COV-2 proteins are highlighted as red diamonds. Differentially methylated genes from the *in vivo* and *in vitro* settings as well as the gene list generated by MCODE are overlapped in the colors yellow, green and blue, respectively. Follow link for a view of the interactive figure.

**Table S1. Demographic characteristics of controls (Con), Covid-19 convalescents (CC19) and symptom-free individuals with T cell responses (SFT).**

|  | **Con** | **CC19** | **SFT** | **Significance** |
| --- | --- | --- | --- | --- |
| ***Female*** | 14/19 (73·7) | 7/14 (50·0) | 4/6 (66·7) | ns |
| ***Age*** | 46.2 (40·8-51·6) | 44·6 (37·4-51·9) | 45·8 (32·4-59·3) | ns |
| ***BMI*** | 23·3 (22·0-24.7)* | 25.3 (22·9-27.7) | 22·8 (20·4-25·3) | ns |
| ***Smoking*** | 2/19 (10·5) | 1/14 (7·1) | 1/5 (20·0)* | ns |

Data from pre-2020 controls (Pre20) is not available, as these samples originate from anonymous blood donors. Comparisons were made using the Pearson χ2 test or Fisher's exact test (<5 observations) for dichotomous variables, or unpaired two-tailed t-tests for the continuous variables. Dichotomous variables are expressed as n/N (%), whereas continuous variables are expressed as mean (95% CI). Pairwise comparisons between the three groups were made, and no significant differences were found. *denotes that data is missing from one individual.

**Table S2. Inferred cell type proportions using the Houseman method from *in vivo* and *in vitro* data.** File supplied separately.

**Table S3, consisting of Tables S3a and S3b.** File supplied separately.

**Table S3a. Differentially methylated CpG sites from analyses on *in vivo* dataset.** A list of differentially methylated CpGs (DMCs) from the comparison between the convalescent Covid-19 subjects and the merged uninfected Con and Pre20 groups. DMCs were defined as CpG sites with a nominal p-value of <0·01 along with a mean methylation difference (MMD) of >0·2.

**Table S3b. Differentially methylated genes from analyses on *in vivo* dataset.** A list of differentially methylated genes (DMGs) from the comparison between the convalescent Covid-19 subjects and the merged uninfected Con and Pre20 groups. DMG were defined as hypermethylated if all DMCs within the gene were hypermethylated and vice versa for hypomethylated genes. If genes contained both hypo- and hypermethylated DMCs, they were named mixed methylation genes.

**Table S4. Significant pathways from over-representation analyses of *in vivo* DMGs.** The PANTHER database was utilised for the analyses and the threshold for significance was set to p<0·05.

|  | **PANTHER Pathway name** | **ref.list** | **Gene count** | **Expected** | **over/under** | **Enrichment** | **p-value** | **Gene ratio** |
| --- | --- | --- | --- | --- | --- | --- | --- | --- |
| **1** | Integrin signalling pathway (P00034) | 193 | 4 | 0·45 | + | 8·89 | 0·00113 | 0·0207253886010363 |
| **2** | Wnt signaling pathway (P00057) | 317 | 3 | 0·74 | + | 4·06 | 0·0382 | 0·00946372239747634 |

| Network module genes (n=66) | |
| --- | --- |
| *ALDH1A2* | *MEF2A* |
| *AP1B1* | *NCOA1* |
| *ARF5* | *NOS3* |
| *ARFIP2* | *NR3C1* |
| *ASAP1* | *NRIP1* |
| *ASAP2* | *NRP1* |
| *CANX* | *NRP2* |
| *CDC37* | *OXT* |
| *CFTR* | *PGR* |
| *CLCN2* | *PGRMC1* |
| *CTSD* | *PHYH* |
| *CYP26B1* | *PLIN3* |
| *CYP51A1* | *PPARA* |
| *DERL1* | *PPARGC1A* |
| *DNAJC5* | *PRKCD* |
| *ESR1* | *PRKCE* |
| *ESRRA* | *PRKG2* |
| *EZR* | *PTPN2* |
| *FAS* | *RASA1* |
| *FKBP4* | *SCNN1A* |
| *GGA1* | *SCNN1G* |
| *GGA2* | *SEMA3F* |
| *GGA3* | *SLC12A2* |
| *GUCY2C* | *SLC26A3* |
| *GZMB* | *SLC2A4* |
| *HDAC6* | *SLC9A3* |
| *HSP90AA1* | *SNAP23* |
| *HSPA4* | *SOX9* |
| *INS* | *SP1* |
| *INS-IGF2* | *SQLE* |
| *KIF13A* | *STX1A* |
| *LAMP1* | *STX8* |
| *M6PR* | *TP53* |

**Table S5. A list of genes from the module identification network analyses using MCODE.** The identified DMGs from the CC19 *vs.* Con+Pre20 comparison (n=54) were used as seed genes in the analyses and applied to the protein-protein interaction network from STRINGdb (confidence score > 0·7), resulting in 66 genes with 139 interactions.

**Table S6. Table of self-reported time between symptoms and sampling in included Covid-19 convalescents.**

| **Sample** | **Weeks between symptoms and sampling** |
| --- | --- |
| CC19_2 | 10-11 weeks |
| CC19_3 | 15-16 weeks |
| CC19_4 | 15-16 weeks |
| CC19_5 | 10 weeks |
| CC19_7 | 15-16 weeks |
| CC19_8 | 18-19 weeks |
| CC19_9 | 15 weeks |
| CC19_10 | 13 weeks |
| CC19_11 | 3-4 weeks |
| CC19_13 | 19-20 weeks |
| CC19_14 | 23-24 weeks |

In the present study, we defined CC19s as individuals having at least SARS-CoV-2-specific IgG antibodies in the circulation (plasma), with possible additional specific immune responses in the saliva and in the T cell compartment as well as the self-reported symptoms. However, three individuals were identified as CC19s by our definition, despite not reporting any symptoms (presumably asympomatic individuals), explaining the discrepancy between the total number of CC19s included in this study (n=14) and the number of individuals present in the above list (n=11).

**Table S7.** A compiled file consisting of analyses performed on the *in vitro* data. Annotated DMC lists from the comparisons of the uninfected pre-2020 blood donors D1-D4 are shown in Tables S7a-c. File supplied separately.

**Table S7a.** A list of annotated CpGs of shared hypermethylated genes (D1-D4, n=1523).

**Table S7b.** Annotated CpG table of shared hypomethylated genes (D1-D4, n=2170).

**Table S7c.** Annotated gene list of shared differentially methylated genes from the in vitro comparison.

**Table S8.** A compiled file consisting of Tables S8a-c, showing overlaps of genes from our analyses that overlap with a publicly available SARS-CoV-2 interactome. File supplied separately.

**Table S8a.** Differentially methylated genes from the in vivo comparison that overlap with the SARS-CoV-2 interactome (n=11)

**Table S8b.** Differentially methylated genes from the in vitro comparison that overlap with the SARS-CoV-2 interactome (n=100)

**Table S8c.** Network module genes from the in vivo comparison that overlap with the SARS-CoV-2 interactome (n=33)
